# Antibiotic self-medication in Otuke district, northern Uganda: Prevalence and associated factors

**DOI:** 10.1101/2024.06.03.24308382

**Authors:** Denis Diko Adoko, Rebecca Nakaziba

**Affiliations:** Department of Midwifery, Lira University, Lira, Uganda; Department of Pharmacology, Lira University, Lira, Uganda

**Author notes:** Corresponding author* / [RN].

**Keywords:** Antibiotic, Self-medication, Prevalence, Factors, Otuke district, Uganda

## Abstract

Antibiotic self-medication is a form of irrational drug use that contributes to antimicrobial resistance, which results in increasing health care costs and morbidity and mortality rates in the population. The misuse of antimicrobial agents is highly linked with the growing problem of antimicrobial resistance within the population globally. Unless addressed, antibiotic self-medication will drive the world back to the pre-antibiotic era, with people dying helplessly due to infectious diseases. This study aimed to investigate the prevalence of antibiotic self-medication and its associated factors in the Otuke District, Northern Uganda. A community-based cross-sectional study was conducted in the Otuke Town Council, Otuke district. The data of adults aged 18 years and above were collected using a semi-structured questionnaire, and the data were coded and entered into SPSS software version 26. The data were descriptively analyzed for frequencies and percentages. Bivariant and multivariant analyses were performed to determine associations between the variables. Out of 385 participants, 68% self-medicated with antibiotics in the past 12 months. Freedom from drug use (AOR: 3.071; 95% CI: 1.203, 7.876) and unregulated use of antibiotics (AOR at 95% CI: 8.288 (2.815, 24.397)) were more likely to lead to ASM (p value <0.001). Other significant factors included knowledge of antibiotics, previous symptom experience, previous successful treatment, long waiting hours and poor staff attitudes (p value <0.05). The most common self-medicated antibiotics were amoxicillin, Ampiclox and metronidazole.

Antibiotic self-medication in the Otuke district is very high due to the availability of medicines and lack of functional drug use regulatory frameworks. The district and government of Uganda should design and implement measures to mitigate this widespread antimicrobial misuse to prevent the development of antimicrobial resistance.

## Introduction

The World Health Organization (WHO) defines self-medication as “the selection and use of medicines by individuals to treat self-recognized illnesses or symptoms without prescription and medical supervision” (1). It is a common form of healthcare practiced in most parts of the world, with over 50% of antibiotics bought and used over the counter (2). Antimicrobial self-medication (ASM) leads to the irrational use of drugs, contributing to antimicrobial resistance, which results in increasing health care costs coupled with increased mortality and morbidity (3, 4). In Africa, the prevalence of ASM ranges from 12.1% to 93.9%, with a mean prevalence of 55.7%. West Africa, as a sub-region, has the highest prevalence at 70.1%, followed by North Africa at 48.1% (5). In Uganda, the prevalence of ASM is as high as 75.7% (Ocan et al., 2014). The misuse of antimicrobial agents is highly linked to the growing problem of antimicrobial resistance (AMR) within the population (6). According to estimates by the World Health Organization (WHO), AMR could lead to 700,000 deaths annually at a global scale, and it has been estimated that 10 million people could die from drug-resistant bacterial infections worldwide by 2050 if the current trend of increasing AMR is not suppressed (7). Antibiotic resistance is a threat to the achievements of modern medicine. Without effective antibiotics to prevent and treat infections, organ transplants, chemotherapy, and surgeries such as cesarean sections are becoming much more dangerous. (8). In developed countries such as Germany, antibiotics are prescription only drugs (9). Unfortunately in developing countries, medicines use, including antibiotics, is rarely regulated (10). In developing countries, antibiotics can be readily purchased without any control, leading to more cases of antibiotic resistance (ABR) (11). The prevalence of ASM in developed countries ranges from 3% to 19%, while that in developing countries ranges from 24% to 73.9% (12). In Africa alone, the prevalence of ASM ranges from 12.1% to 93.9%, with a mean prevalence of 55.7% (5). For instance, in a studies conducted in Eritrea, Ghana, and Sudan; the prevalence of ASM was 45.1%, 36%, 71.3% respectively (13-15). The main determinants of ASM include education level, age, gender, past successful use, severity of illness and income (16). According to Green, Keenan (17) whereas in low- and middle-income countries, past good experiences and suggestions from friends or relatives promote self-medication (18). In Eritrea, the main reasons for ASM were also previous successful experience and the illness being ‘not serious enough to seek medical care’ (13) but in Afghanistan, economic problems, lack of time to visit doctors, and ease of use were cited as the main reasons for practicing ASM (19). In Pakistan, ASM is affected by male gender, age, and education level (20). In a related study performed in Moshi, Kilimanjaro, among university students, shortage of drugs at health facilities, long waiting times at health facilities and long distances to health facilities were the main causes of ASM (12). In a similar study, inability to pay for health care charges, the freedom to choose the preferred drugs, lack of medical professionals, poor quality of healthcare facilities, unregulated distribution of medicines and patients’ misconception about physicians were factors for ASM in developing countries (21). According to a study performed in Afghanistan the commonly self-medicated antibiotics were penicillins, metronidazole, and ceftriaxone (19) while in Pakistan, Ciprofloxacin was the most commonly used self-medicated (20). In Western Uganda, Amoxicillin and metronidazole are the most commonly used antibiotics among nursing students (22) whereas in northern Uganda, cotrimoxazole, amoxicillin and metronidazole are the most commonly used (23).

Self-medication with antibiotics results in health problems such as treatment failure, an increased incidence of side effects and the development of acquired AMR, which decreases the effectiveness of antibiotics for treating bacterial infections and is now becoming a global health problem (10). The development of antibiotics was one of the great discoveries of modern medicine (24). Antibiotics fight bacteria and can cure life-threatening infectious diseases such as pneumonia, for which there was previously no effective treatment (21). However, the improper use of antibiotics has led to an increasing number of bacteria becoming resistant to these types of drugs (25). Antimicrobial resistance (AMR) is currently increasing. It has been a global public health concern for the past decade (26). The number of antibiotic-resistant microorganisms is on the rise (27), making it difficult to achieve good treatment outcomes during the management of infectious diseases. The costs of AMR to economies and their healthcare systems are significant, affecting the productivity of patients and their caregivers through prolonged hospital stays and the need for more expensive and intensive care (28). Although AMR is a natural phenomenon, resistance develops more rapidly through the misuse and overuse of antimicrobial agents (28, 29). Therefore, the public plays a crucial role in the development or prevention of this disease. The current study aimed to determine the prevalence of antibiotic self-medication in Otuke District, Northern Uganda, and the associated factors to inform the government of the vice so that informed interventions can be designed.

## Materials and methods

### Study Area

The study was performed in the Otuke District. Otuke District is located within the Lango subregion in Northern Uganda. Otuke district consists of many government health facilities that are well distributed across all sub-counties and a total of 50 drug shops. The Otuke Town Council alone has five medical clinics (*Source: Records from the National Drug Authority, Otuke District)*.

### Participant recruitment and procedures

The study was a descriptive cross-sectional study employing quantitative methods of data collection. A cross-sectional design is time-saving and inexpensive. The sample size was calculated using the Kish Leslie (1965) formula assuming a proportion of 50% and yielded a sample size of 385. The study was conducted among adult community members (aged 18 years and above) of the Otuke District excluding those who were very sick and could not respond to the questions. The study employed a cluster sampling technique. The clusters were randomly selected from the Adwon-Ibuto, Barodugu, Teogini and Te-Boke cells of the Otuke Town Council. The study participants were conveniently selected from each cluster and the potential participants were accessed by the research team moving into their homes. The study purpose, risks and benefits were clearly explained to potential participants who were required to sign an informed consent form to affirm their acceptance. Those who agreed to participate in the study and signed the consent form were recruited. The data were collected using a pretested questionnaire developed in line with the study objectives. The tool had three parts: Part A (Sociodemographic characteristics such as age, sex, and level of education), Part B (Self-medication practices), and Part C (Factors associated with ASM (Health system factors, individual factors, and interpersonal factors). Following the acquisition of informed consent, data were collected using an interviewer-administered questionnaire. Each section of the questionnaire was well explained to the respondents who then provided answers. The questionnaires were checked for completeness after data collection, and those with incomplete or irrelevant data were excluded from the analysis. Following data collection, the data were coded and double-entered into Statistical Package for Social Sciences (SPSS) software version 26 and descriptively analyzed for frequencies and percentages. Bivariate and multivariate analyses were performed to identify associations between the variables.

#### Ethical issues

The study was presented before the faculty of Nursing and Midwifery, Lira University and to Otuke district administration for approval. Moreover, the study purpose, risks and benefits were clearly explained to potential participants (adults aged 18 years and above) who were required to sign an informed consent form to affirm their acceptance before recruitment into the study.

## Results

### Socio-demographic data

A total of 49.4% of the participants were aged between 18 and 28 years, 56.6% were females, 54.3% stopped at the primary level of education, 53.5% were married, 65.5% were self-employed, 94.8% were members of the Lango tribe, and 44.9% were Anglicans (Table 1).

**Table 1:** Socio-demographic characteristics of the study participants (n=385)

| Variable | Frequency(f) | Percentage (%) |
| --- | --- | --- |
| **Age** |  |  |
| 18-28 | 190 | 49.4 |
| 29-40 | 114 | 29.6 |
| 41-60 | 64 | 16.6 |
| 61 and above | 17 | 4.4 |
| **Gender** |  |  |
| Male | 167 | 43.4 |
| Female | 218 | 56.6 |
| **Level of education** |  |  |
| Primary | 209 | 54.3 |
| Secondary | 121 | 31.4 |
| Tertiary | 47 | 12.2 |
| No education | 8 | 2.1 |
| **Marital status** |  |  |
| Single | 153 | 39.7 |
| Married | 206 | 53.5 |
| Separated | 26 | 6.8 |
| **Occupation** |  |  |
| Self-employed | 252 | 65.5 |
| Government employee | 40 | 10.4 |
| Employed in the private sector | 30 | 7.8 |
| Not employed | 63 | 16.4 |
| **Tribe** |  |  |
| Lango | 365 | 94.8 |
| Others | 20 | 5.2 |
| **Religion** |  |  |
| Anglican | 173 | 44.9 |
| Seventh-Day Adventist | 16 | 4.2 |
| Islam | 8 | 2.1 |
| Catholic | 155 | 40.3 |
| Pentecostal | 31 | 8.1 |
| Others | 2 | 0.5 |

### Prevalence of antibiotic self-medication

The majority of the community members of the Otuke Town Council (68%) practiced self-medication (Fig 1)

**Fig. 1:**
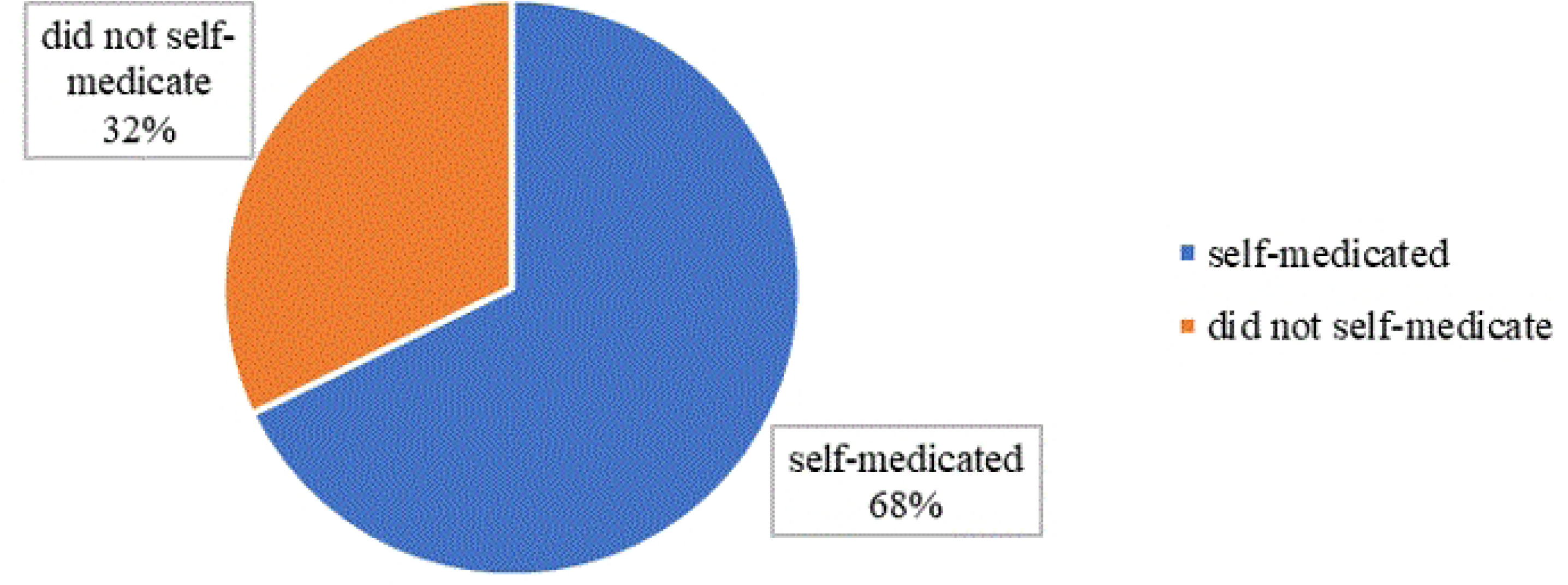
Prevalence of antibiotic self-medication in Otuke district (n=385).

### Commonly self-medicated antibiotics and their sources

The most common self-medicated antibiotic was amoxicillin (51.3%). Most of the respondents (98.5%) obtained antibiotics from drug shops/pharmacy (Table 2).

**Table 2:**
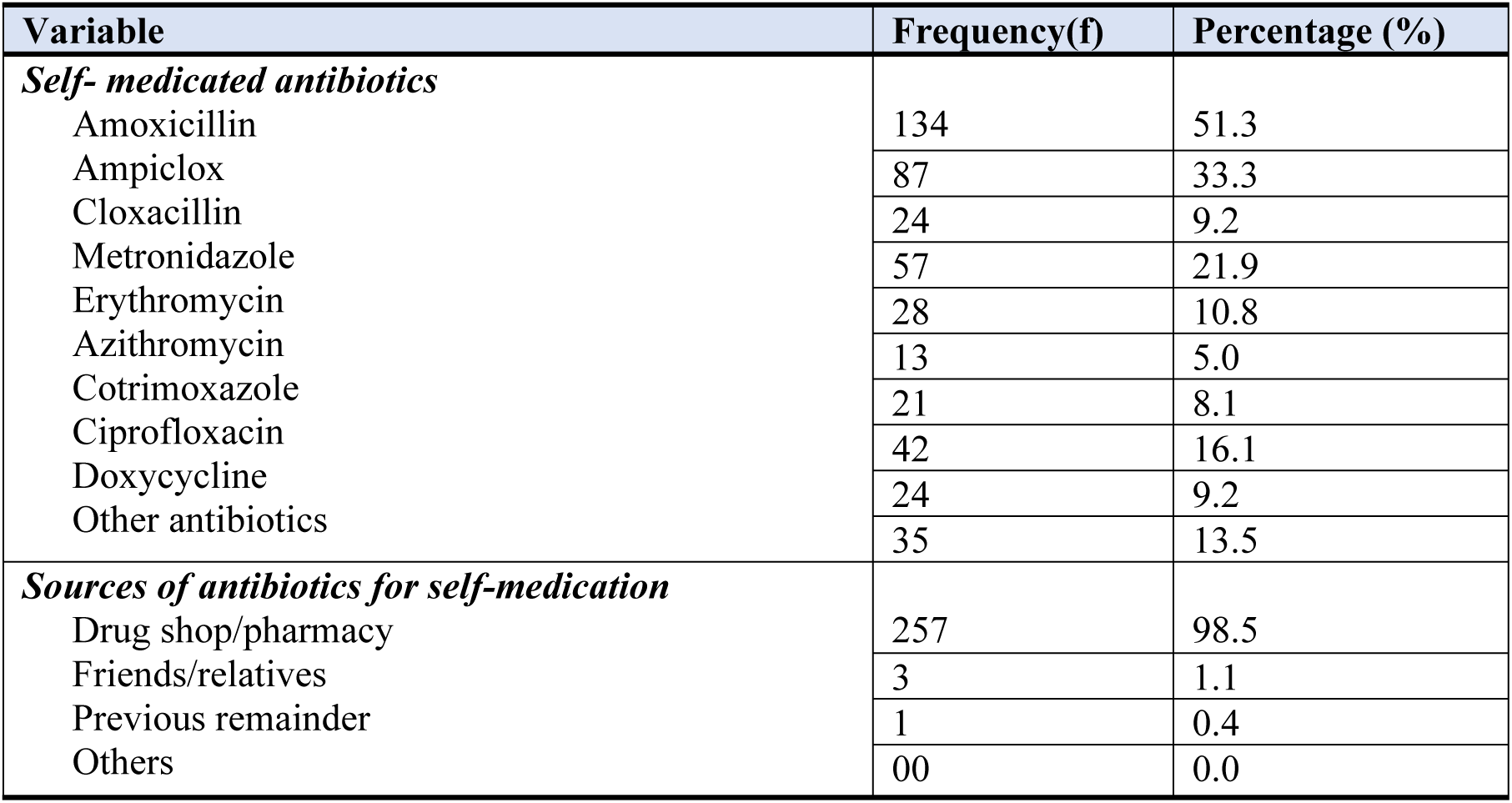
Commonly self-medicated antibiotics and their sources (n=385)

| Variable | Frequency(f) | Percentage (%) |
| --- | --- | --- |
| ***Self- medicated antibiotics*** |  |  |
| Amoxicillin | 134 | 51.3 |
| Ampiclox | 87 | 33.3 |
| Cloxacillin | 24 | 9.2 |
| Metronidazole | 57 | 21.9 |
| Erythromycin | 28 | 10.8 |
| Azithromycin | 13 | 5.0 |
| Cotrimoxazole | 21 | 8.1 |
| Ciprofloxacin | 42 | 16.1 |
| Doxycycline | 24 | 9.2 |
| Other antibiotics | 35 | 13.5 |
| ***Sources of antibiotics for self-medication*** |  |  |
| Drug shop/pharmacy | 257 | 98.5 |
| Friends/relatives | 3 | 1.1 |
| Previous remainder | 1 | 0.4 |
| Others | 00 | 0.0 |

### Factors associated with Antibiotic self-medication

#### Health system, individual and interpersonal factors

Among the health system factors, individual factors and interpersonal factors; long waiting hours (82.9%), previous successful treatment (55.6%), and suggestions by friends/relatives (24.4%) had the greatest effects respectively (Table 3).

**Table 3:** Health system, individual and interpersonal factors associated with ASM.

| Variable | Frequency(f) | Percentage (%) |
| --- | --- | --- |
| **Health system factors** |  |  |
| Poor staff attitude | 110 | 28.6 |
| Health facility too far | 55 | 14.3 |
| Long waiting hours | 319 | 82.9 |
| No drugs at the health facility | 268 | 69.9 |
| Lack of medical professional | 35 | 9.1 |
| Unregulated distribution of antibiotics | 89 | 23.1 |
| **Individual factors** |  |  |
| Previous symptom experience | 207 | 53.8 |
| Previous successful treatment | 214 | 55.6 |
| Illness was not serious, | 211 | 54.8 |
| Storage of antibiotics | 109 | 28.3 |
| Lack of time to visit doctors | 70 | 18.2 |
| Misconception about health workers | 21 | 5.5 |
| Inability to pay for the health charges | 48 | 12.5 |
| Freedom to choose the drugs | 177 | 46.1 |
| Knowledge on the antibiotics | 157 | 40.8 |
| Old prescription | 113 | 29.4 |
| **Interpersonal factors** |  |  |
| Given by a friend | 43 | 11.2 |
| Suggestion by a relative/friend | 94 | 24.4 |
| Drug brought by health worker | 41 | 10.6 |

### Associations between socio-demographic characteristics with ASM

There was no significant association between socio-demographic characteristics and antibiotic self-medication in the present study (Table 4).

**Table 4:** Association of sociodemographic characteristics with ASM (at 95% CI)

| Variables | Antibiotic Self-medication: Freq (%) |  | X<sup>2</sup> | df | p value |
| --- | --- | --- | --- | --- | --- |
|  | Yes | No |  |  |  |
| Age |  |  |  |  |  |
| 18-28 | 127(48.7) | 63(50.8) |  |  |  |
| 29-40 | 80(30.7) | 34(27.4) | 1.369 | 3 | 0.694 |
| 41-60 | 41(15.7) | 23(18.5) |  |  |  |
| 61 and above | 13(5.0) | 4(3.2) |  |  |  |
| Gender |  |  |  |  |  |
| Male | 114(43.7) | 53(42.7) | 0.030 | 1 | 0.862 |
| Female | 147(56.6) | 71(57.3) |  |  |  |
| **Level of education** |  |  |  |  |  |
| Primary | 137(52.5) | 72(58.1) |  |  |  |
| Secondary | 78(29.9) | 43(34.7) | 7.483 | 3 | 0.862 |
| Tertiary | 39(14.9) | 8(6.5) |  |  |  |
| No education | 7(2.7) | 1(0.8) |  |  |  |
| **Marital status** |  |  |  |  |  |
| Single | 107(40.2) | 48(38.7) |  |  |  |
| Married | 140(53.6) | 66(53.2) | 0.517 | 2 | 0.772 |
| Separated | 16(6.1) | 10(8.1) |  |  |  |
| **Occupation** |  |  |  |  |  |
| Self-employed | 164(62.8) | 88(71.0) |  |  |  |
| Government employee | 30(11.5) | 10(8.1) | 2.655 | 3 | 0.448 |
| Employed in the private sector | 21(8.0) | 9(7.3) |  |  |  |
| Not employed | 46(17.6) | 17(13.7) |  |  |  |
| **Tribe** |  |  |  |  |  |
| Lango | 246(94.3) | 119(96.0) | 0.502 | 1 | 0.479 |
| Others | 15(5.7) | 5(4.0) |  |  |  |
| **Religion** |  |  |  |  |  |
| Anglican | 121(69.9) | 52(30.1) |  |  |  |
| Seventh-Day Adventist | 12(75.0) | 4(25.0) |  |  |  |
| Islam | 6(75.0) | 2(25.0) | 3.511 | 5 | 0.622 |
| Catholic | 102(65.8) | 53(34.2) |  |  |  |
| Pentecostal | 18(58.1) | 13(41.9) |  |  |  |
| Others | 2(0.5) | 0(100) |  |  |  |

### Associations between health system factors with ASM

Most health system factors were strongly associated with antibiotic self-medication (p value <0.001) (Table 5).

**Table 5:**
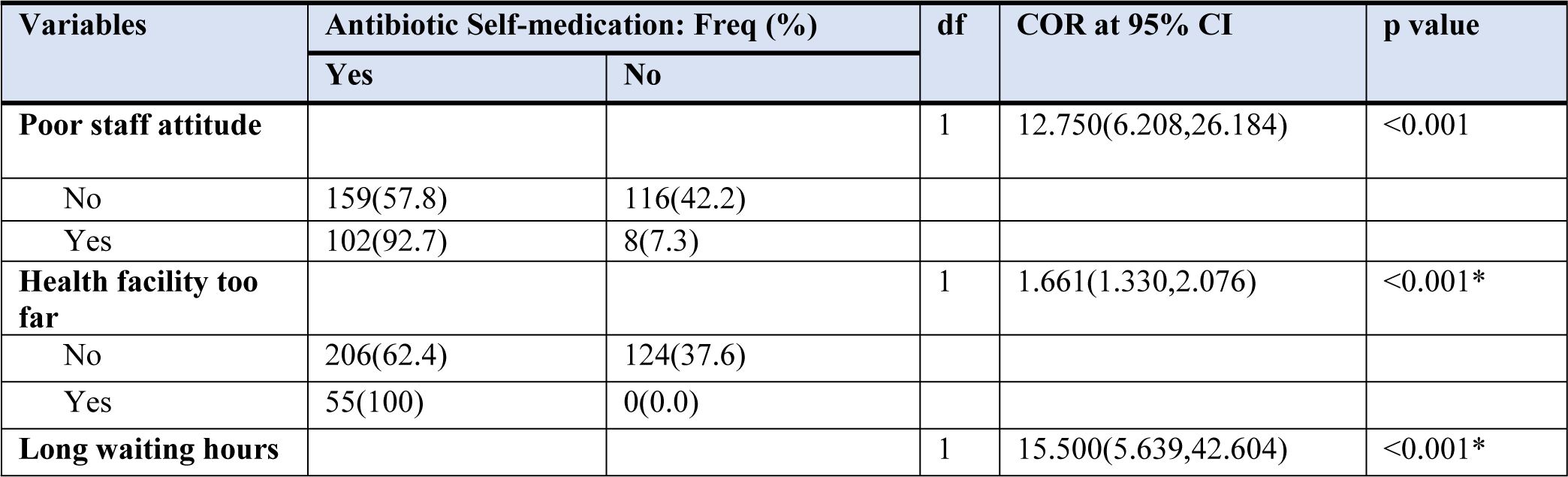
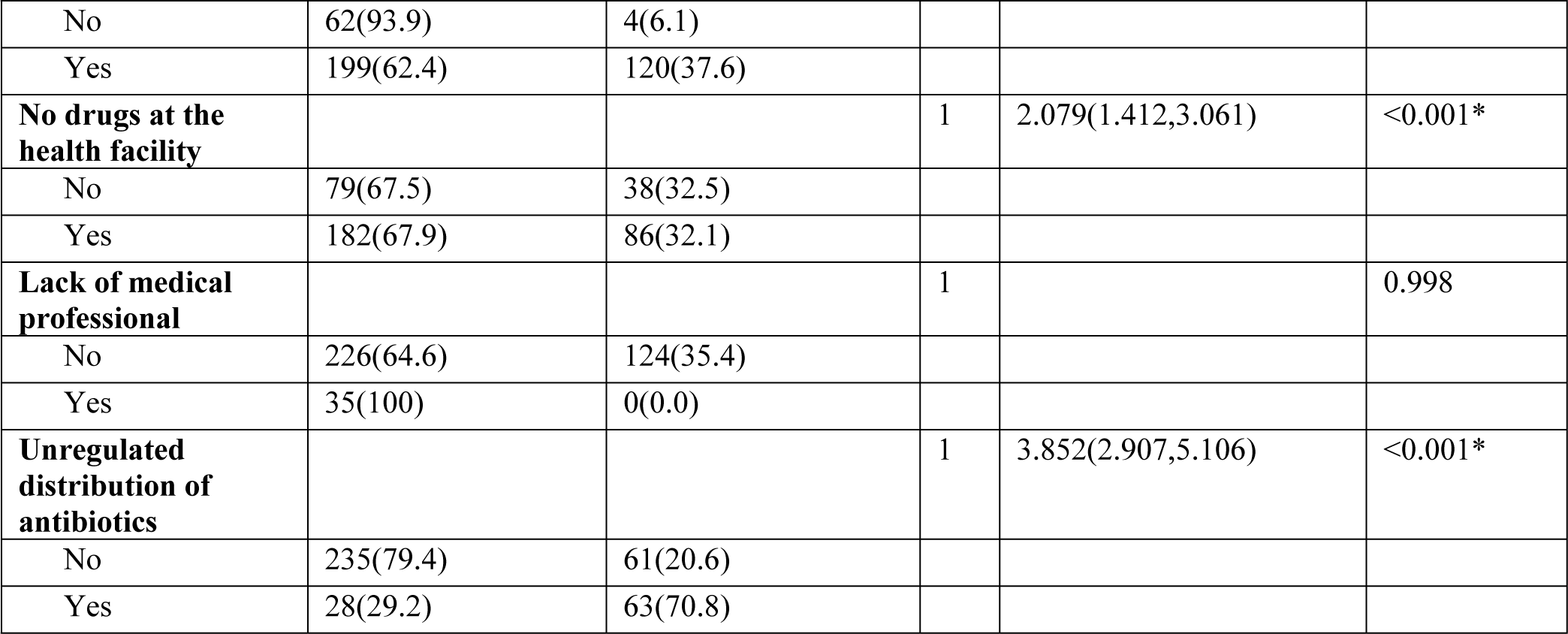
Association of health system factors with ASM.

| Variables | Antibiotic Self-medication: Freq (%) |  | df | COR at 95% CI | p value |
| --- | --- | --- | --- | --- | --- |
|  | Yes | No |  |  |  |
| **Poor staff attitude** |  |  | 1 | 12.750(6.208,26.184) | <0.001 |
| No | 159(57.8) | 116(42.2) |  |  |  |
| Yes | 102(92.7) | 8(7.3) |  |  |  |
| **Health facility too far** |  |  | 1 | 1.661(1.330,2.076) | <0.001* |
| No | 206(62.4) | 124(37.6) |  |  |  |
| Yes | 55(100) | 0(0.0) |  |  |  |
| **Long waiting hours** |  |  | 1 | 15.500(5.639,42.604) | <0.001* |
| No | 62(93.9) | 4(6.1) |  |  |  |
| Yes | 199(62.4) | 120(37.6) |  |  |  |
| **No drugs at the health facility** |  |  | 1 | 2.079(1.412,3.061) | <0.001* |
| No | 79(67.5) | 38(32.5) |  |  |  |
| Yes | 182(67.9) | 86(32.1) |  |  |  |
| **Lack of medical professional** |  |  | 1 |  | 0.998 |
| No | 226(64.6) | 124(35.4) |  |  |  |
| Yes | 35(100) | 0(0.0) |  |  |  |
| **Unregulated distribution of antibiotics** |  |  | 1 | 3.852(2.907,5.106) | <0.001* |
| No | 235(79.4) | 61(20.6) |  |  |  |
| Yes | 28(29.2) | 63(70.8) |  |  |  |

### Associations between interpersonal factors with ASM

There was a very strong association between most interpersonal factors and self-medication (p value<0.05) (Table 6).

**Table 6:** Association between interpersonal factors with ASM.

| Variables | Antibiotic Self-medication: Freq (%) |  | df | COR at 95% CI | p value |
| --- | --- | --- | --- | --- | --- |
|  | No | Yes |  |  |  |
| **Given by a friend** |  |  | 1 | 1.758(1.410,2.192) | <0.001* |
| No | 124(36.3) | 218(63.7) |  |  |  |
| Yes | 0(0.0) | 43(100) |  |  |  |
| **Suggestion by a relative/friend** |  |  | 1 | 93.000(12.963,667.189) | <0.001* |
| No | 123(42.3) | 168(57.7) |  |  |  |
| Yes | 1(1.1) | 93(98.9) |  |  |  |
| **Drug brought by health worker** |  |  | 1 | 0.640(0.591,0.692) | 0.997 |
| No | 124(36.0) | 220(64.0) |  |  |  |
| Yes | 0(0.0) | 41(100) |  |  |  |

### Associations of individual factors with ASM

As indicated in Table 7 below, there was a strong association between most of the individual factors and ASM (p value <0.001).

**Table 7:** Association of individual factors with ASM.

| Variables | Antibiotic Self-medication: Freq (%) |  | df | COR at 95% CI | p value |
| --- | --- | --- | --- | --- | --- |
|  | No | Yes |  |  |  |
| Previous symptom experience |  |  | 1 | 3.684(2.574,5.273) | <0.001* |
| No | 38(21.3) | 140(78.7) |  |  |  |
| Yes | 86(41.5) | 121(58.5) |  |  |  |
| Previous successful treatment |  |  | 1 | 0.462(0.334,0.637) | <0.001* |
| No | 117(68.4) | 54(31.6) |  |  |  |
| Yes | 7(3.3) | 207(96.7) |  |  |  |
| Illness was not serious |  |  | 1 | 0.813(0.603,1.095) | 0.173 |
| No | 96(55.5) | 78(44.8) |  |  |  |
| Yes | 28(13.3) | 183(86.7) |  |  |  |
| Storage of antibiotics |  |  | 1 | 1.226(0.967,1.554) | 0.092 |
| No | 124(44.9) | 152(55.1) |  |  |  |
| Yes | 0(0.00) | 109(100) |  |  |  |
| Lack of time to visit doctors |  |  | 1 | 1.540(1.229,1.931) | <0.001* |
| No | 124(39.4) | 191(60.6) |  |  |  |
| Yes | 0(0.00) | 70(100) |  |  |  |
| Misconception about health workers |  |  | 1 | 1.935(1.558,2.404) | <0.001* |
| No | 124(34.1) | 240(65.9) |  |  |  |
| Yes | 0(0.0) | 21(100) |  |  |  |
| Inability to pay for the health charges |  |  | 1 |  | 0.997 |
| No | 124(36.8) | 213(63.2) |  |  |  |
| Yes | 0(0.0) | 48(100) |  |  |  |
| Freedom to choose the drugs |  |  | 1 | 0.580(0.428,0.788) | <0.001* |
| No | 12(5.8) | 195(94.2) |  |  |  |
| Yes | 112(63.3) | 65(36.7) |  |  |  |
| Knowledge on the antibiotics |  |  | 1 | 5.162(3.630,7.340) | <0.001* |
| No | 37(16.2) | 191(83.8) |  |  |  |
| Yes | 87(55.4) | 70(44.6) |  |  |  |
| Old prescription |  |  | 1 |  | 0.996 |
| No | 124(45.6) | 148(54.4) |  |  |  |
| Yes | 0(0.0) | 113(100) |  |  |  |

**Legend:** CI- confidence interval \* significant p value

### Multivariate analysis of the association of factors with ASM

The results from multivariate analysis of variables showed that freedom to choose drugs (AOR: 3.071; 95% CI (1.203, 7.876)) and unregulated use of antibiotics (AOR at 95% CI: 8.288 (2.815, 24.397)) were more likely to lead to ASM (p value <0.001). Other significant factors included knowledge of antibiotics, previous symptom experience, previous successful treatment, long waiting hours and poor staff attitudes (p value <0.05) (Table 8).

**Table 8:**
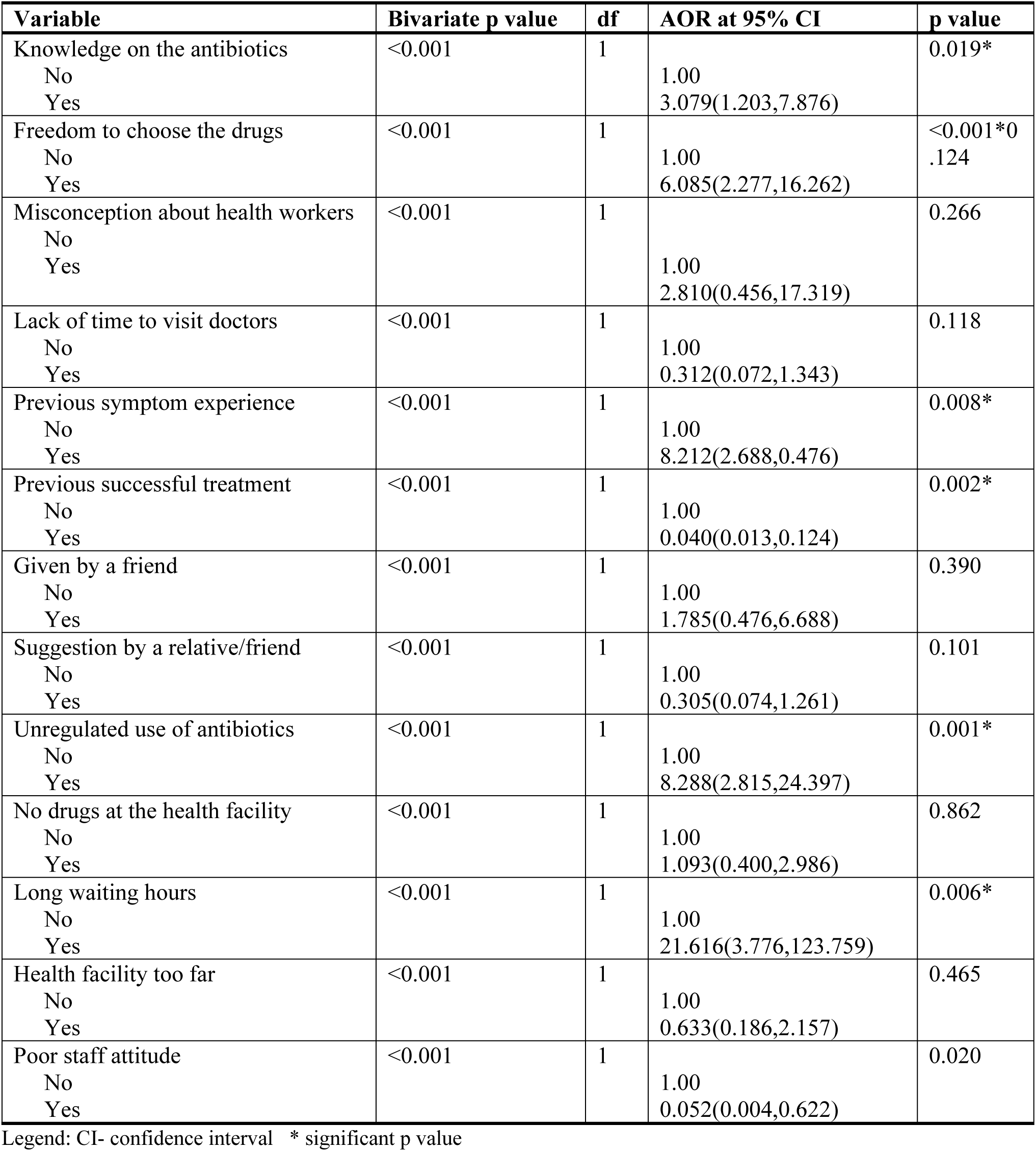
Association between factors and antibiotic self-medication.

## Discussion

ASM is one of the most common causes of AMR worldwide; however, it can be controlled if appropriate measures are taken. The ability to develop preventive measures can only be made possible by tracking the self-medication behaviors of people in the public. Additionally, it reveals the health system’s weaker areas. To ascertain the prevalence of ASM in the Otuke District and to gain insight into the contributing factors of ASM, the current cross-sectional study was carried out.

Of the 385 participants, 68% (over two-thirds) reported having self-medicated with antibiotics in the past 6 months. This implies that approximately 7 in 10 community members practiced ASM, with 49.4% of ASM being in the age bracket 18-28 years. This could be attributed to the fact that there are no regulations in place to curb the practice, so every member of the community is free to buy antibiotics from the many established drug shops or pharmacies. This could also be the result of the long waiting hours experienced when patients seek health care services in various health facilities. The findings of this study are closely related to those of a similar study in Sudan, where the prevalence of ASM was 71% (15); and to another in northern Uganda, which was 73% (23); and in Afghanistan, at 73.2% (19). Against this background, a way forward is worth seeking to reduce the practice of ASM among the community. On the other hand, a similar study conducted among nursing students in Western Uganda showed a much greater prevalence of ASM, at 85.7% (22). This could be due to the pharmacological knowledge gained by the nursing students. In contrast, related studies in the Middle East and South America revealed lower prevalence rates of ASM, at 36.1% to 45.8% and 29%, respectively (12). This could be due to better drug use regulations that guide antimicrobial use in these countries, whereby antibiotics are prescription only drugs.

A variety of factors were analyzed for their association with ASM in this study grouped into health system, individual and interpersonal factors. The findings indicated that freedom to choose drugs (AOR: 3.071; 95% CI (1.203, 7.876)) and unregulated use of antibiotics (AOR at 95% CI: 8.288 (2.815, 24.397)) were more likely to lead to ASM (p value <0.001). This is true because in Uganda, the use of antibiotics is not highly regulated, so members of the public have free access to these antibiotics. This finding is similar to that of Sachdev and others who found that unregulated deliveries were the major driver of ASM (30). Therefore, the enactment of drug regulation laws could be the beginning of an end to ASM. Previous symptom experience was also significantly associated with ASM probably because individuals tend to associate a given symptom with a disease. Therefore, a symptom managed successfully in the past is deemed manageable in the same way in the future. This is in agreement with findings of a research conducted in Pakistan (31). Although many symptoms may be successfully managed by a specific drug, different conditions can manifest in a similar way, leading to the irrational use of antibiotics. Communities should be aware that some diseases mimic each other and that the use of a given antibiotic could be more of a disaster than a blessing. In addition, previous successful treatment was highly significantly associated with ASM. This could have been due to the patient’s ability to remember how he/she was managed. This finding is consistent with a study performed in low- and middle-income countries (LMICs) in which the habit of self-medication using previous treatments was identified (18). There is a need for more patient and family sensitization to the dangers of practicing ASM. Furthermore, long waiting hours in health facilities were reported by multiple respondents in the present study were associated with ASM. Many respondents thought it was a waste of time going to a health facility at 8:00 am and returning home late in the afternoon. Therefore, they end up leaving the facility in search of quicker services at drug stores. In Ethiopia, a similar study supported the present findings where long waiting times contributed to self-medication (32). Increased staffing could reduce the time spent at the facility during health seeking and improve patient trust. Moreover, poor staff attitude was another factor that was significantly associated with ASM in this study. Staff attitudes in terms of being rude, asking for money or not willing to help caused the community members to go elsewhere and buy their own drugs without consultation. This finding was consistent with those from studies in Cameroon and LMICs (33, 34). Regular staff motivation and appraisal could instill and sustain the drive to serve the medical staff, which could improve medication practices with antibiotics and prevent ASM.

Of the 385 participants, 98.5% bought antibiotics from drug shops/pharmacies. This study also explored the common self-medicated antibiotics and discovered Amoxicillin (51.3%), Ampiclox (33.3%), and Metronidazole(21.9%). In western Uganda among nursing students, Amoxicillin and Metronidazole were the most commonly used antibiotics (22) as well as in northern Uganda as reported by Ocan, Bwanga (23). In Sudan, Amoxicillin is the most commonly used antibiotic (15). The increased use of Amoxicillin could be due to its lower cost than other antibiotics and the high prevalence of respiratory tract infections (although not all respiratory infections require antibiotics for treatment) (35). Sensitizing the community to various treatment modalities for respiratory infections could be vital for minimizing ASM.

## Conclusion and recommendations

Self-medication with antibiotics is highly practiced in the Otuke District. Factors leading to ASM originate from the health system as well as from individuals predisposing the community to the danger of antimicrobial resistance if not addressed.

The health team should provide adequate information regarding the dangers of ASM to the general public and equip community members with knowledge regarding supportive ways of managing common conditions. In addition, the government should enact drug laws that govern the use of antimicrobial agents and improve staff welfare to motivate them to effectively deliver their services to the community. Furthermore, the government should recruit more staff to quicken health service delivery.

## Data Availability

The data are available from the corresponding author upon reasonable request or at https://ir.lirauni.ac.ug/xmlui/

## Acknowledgments

The community of the Otuke district for participating in this study.

## Author contributions

ADD: Conception, methods, data collection; reporting and review; RN: Methods, supervision, manuscript draft and review

## Notes

### Competing Interest Statement

The authors have declared no competing interest.

### Funding Statement

The author(s) received no specific funding for this work.

### Author Declarations

Lira University, Faculty of Nursing and Midwifery, Research Ethics Committee.

